# Direct RNA nanopore sequencing of SARS-CoV-2 extracted from critical material from swabs

**DOI:** 10.1101/2020.12.21.20191346

**Authors:** Davide Vacca, Antonino Fiannaca, Fabio Tramuto, Valeria Cancila, Laura La Paglia, Walter Mazzucco, Alessandro Gulino, Massimo La Rosa, Carmelo Massimo Maida, Gaia Morello, Beatrice Belmonte, Alessandra Casuccio, Rosario Maugeri, Gerardo Iacopino, Francesco Vitale, Claudio Tripodo, Alfonso Urso

**Affiliations:** University of Palermo, Department of Sciences for Health Promotion and Mother-Child Care “G. D’Alessandro”, Piazza delle cliniche N°2, zip code 90127, Palermo, Italy; CNR-ICAR, National Research Council of Italy, Via Ugo La Malfa, a5c, Palermo, Italy; Cogentech srl Società Benefit, FIRC Institute of Molecular Oncology (IFOM) Via Adamello 16, 20139, Milan, Italy; Department of Experimental Biomedicine and Clinical Neurosciences, School of Medicine, Neurosurgical Clinic, University of Palermo, Palermo, Italy

**Keywords:** MinION, Direct RNA Nanopore sequencing, Sars-CoV-2, Covid19, Swab

## Abstract

**Background:** In consideration of the increasing prevalence of COVID-19 cases in several countries and the resulting demand for unbiased sequencing approaches, we performed a direct RNA sequencing experiment using critical oropharyngeal swab samples collected from Italian patients infected with SARS-CoV-2 from the Palermo region in Sicily.

**Methods:** Here, we identified the sequences SARS-CoV-2 directly in RNA extracted from critical samples using the Oxford Nanopore MinION technology without prior cDNA retro-transcription.

**Results:** Using an appropriate bioinformatics pipeline, we could identify mutations in the nucleocapisid (N) gene, which have been reported previously in studies conducted in other countries.

**Conclusion:** To the best of our knowledge, the technique used in this study has not been used for SARS-CoV-2 detection previously owing to the difficulties in the extraction of RNA of sufficient quantity and quality from routine oropharyngeal swabs.

Despite these limitations, this approach provides the advantages of true native RNA sequencing, and does not include amplification steps that could introduce systematic errors.

This study can provide novel information relevant to the current strategies adopted in SARS-CoV-2 next-generation sequencing.

We deposited the gene sequence in the NCBI database under the following URL:https://www.ncbi.nlm.nih.gov/nuccore/MT457389

## INTRODUCTION

The study of the SARS-CoV-2 genome has become a priority for global healthcare to facilitate the identification of more suitable vaccines and therapeutic drugs (1). The use of third generation sequencing technologies has increased significantly in recent years, as these methods yield reliably long reads even from biological samples with noise (2).

Direct RNA sequencing (RNA-seq) of SARS-CoV-2 has the advantage of displaying its native sequence without contamination by artefacts originating from the in vitro cDNA amplification step, which is often necessary for target enrichment (3). However, the quality of RNA extracted from the swab has certain critical issues with respect to both fragmentation level and concentration, such that the read coverage of sequencing decreases considerably. In this study, we investigate the suitability of Nanopore Oxford MinION Mk1B (4), a third-generation nanopore-based platform, for the identification of a critical target of SARS-CoV-2 native RNA from a swab sample, and its sensitivity in the characterization of the viral mutational landscape.

To this end, we analyzed ten RNA samples extracted from oropharyngeal swabs of ten patients with COVID-19, and sequenced them in two pools following the direct RNA sequencing protocol (SQK-RNA002, Nanopore Technologies). Although the concentration of the library loaded in the sequencing flow cell was approximately 200 times lower than that required for the protocol, we clearly identified two mutations in the nucleocapsid phosphoprotein (N) gene with respect to the sequence of the strain isolated in Wuhan, which has been described in literature.

## MATERIALS AND METHODS

After the collection of ten oropharyngeal swab samples from Italian patients with COVID-19, we assayed the concentration and fragmentation level of the extracted RNA to arrange it for sequencing, as recommended by Oxford Nanopore in the Input DNA/RNA quality control guidelines (5). Next, we prepared two sequencing libraries from two distinct pools, as described below, and launched a computational pipeline for aligning the obtained reads. Lastly, we confirmed the mutation frequency in our sample either by Sanger sequencing or by real-time PCR (qPCR) assays.

### Input RNA collection and quality control steps

#### Samples

For this study, we collected RNA samples from ten Sicilian patients between March 19^th^ and 23^rd^, 2020. The patients tested positive in the 2019-Novel Coronavirus (2019-nCoV) Real-time rRT-PCR Panel (Centers for Disease Control and Prevention (CDC) Atlanta, GA 30333).

Furthermore, to compare the nanopore sequencing results, we included two samples from uninfected individuals as negative controls. The RNA from uninfected controls was extracted from brain biopsy specimens of two patients who underwent surgery in August 2019 and in January 2020, respectively. Both biopsies were made available from Department of Experimental Biomedicine and Clinical Neurosciences, School of Medicine, Neurosurgical Clinic, University of Palermo.

#### RNA extraction

The swab buffer kit was assayed using the QIAamp Viral RNA Mini Kit in Automated purification of RNA on QIAcube Instruments (Qiagen, Cat.No 9001292) according to the manufacturer’s instructions.

#### Assessment of concentration and fragmentation level

The quality of the fragmented sample was assessed with an Agilent Bioanalyzer 2100 (Agilent, Cat.No G2939BA) using the Agilent RNA 6000 Pico assay (Agilent, Cat.No 5067-1513). We assayed 1 µL of each pure sample. The RNA extracted from the samples showed high fragmentation and lowest concentration levels with an RIN index between 2.6 and 2.1 and concentration between 19 and 829 pg/µL.

### Preparation of the libraries and the computational pipeline

The samples were organized in two pools (A and B), each with a final volume of 10 µL. Pool A included samples from three patients: two with a PCR cycle threshold (Ct) value of 18 and one with Ct value of 20. To elaborate, we pooled 8 µL of samples from the first two patients (4 µL from each) and 2 µL from the last patient.

Pool B included RNA samples collected from ten patients (1 µL from each patient): three samples were from pool A, while the other seven had decreasing Ct values, and were collected from two patients with Ct value of 21, two with 22, two with 23, and one with 24.

#### Library preparation, priming, and commencement of a sequencing run

Both pools were sequenced using a MinION Mk1B sequencer with the SQK-RNA002 protocol. After preparing the sequencing libraries A and B, as previously described, to ligate retro-transcription adapters (RTA) to the 3’-end of the RNA molecules, we combined each library with 6 µL of a mix comprising 3 µL of NebNext Quick Ligation Reaction Buffer, 1.5 µL of T4 DNA Ligase, 1 µL of RTA, and 0.5 µL of 110 nM RNA CS (RCS). Each reaction was incubated for 10 min at 22 °C. Next, the reactants were mixed with 9 µL of nuclease-free water, 8 µL of 5X first-strand buffer, 4 µL of 0.1 DTT, and 2 µL of 10 mM dNTPs. For cDNA synthesis, we added 2 µL of SuperScript III, following which both reactions were incubated under the following conditions: 50 °C for 50 min, 70 °C for 10 min, and 4 °C before proceeding to the next step.

The subsequent wash step was performed according to the procedure for the specified protocol.

Next, we measured the concentration of 1 µL of each library using a Qubit 3.0 Fluorimeter and the dsDNA HS assay kit (Life Technologies, Cat. No Q32851). The concentrations were 1.2 ng/µL and 0.6 ng/µL for libraries A and B, respectively.

Next, we performed the Attachment of 1D sequencing adapter step according to manufacturer instructions, and recorded final concentrations of 0.9 ng/µL and 0.3 ng/µL for A and B, respectively.

Lastly, 20 µL of sample from each library was mixed with 17.5 µL of nuclease-free water and 37.5 µL of RNA running buffer (RRB) in a final volume of 75 µL.

The assessment of the R9.4 Flow Cell, which was performed prior to the loading of the libraries, revealed that 1575 active pores were available for sequencing.

We performed a new MinKNOW (6) (*v19*.*12*.*5*) experiment using only the A loading mix in the flow cell.

In MinKNOW we selected the SQK-RNA002 Kit and continued the sequencing process until approximately 80% pores were available (i.e., after 4 h). At this point, we paused the process and loaded the B loading mix in the flow cell. Next, the run was resumed and continued till the number of reads generated was unvaried at 397k (i.e., after approximately 27 h). At the end of the sequencing process, we obtained the “Fast5” raw data files.

#### Computational pipeline

We developed a computational pipeline to analyze the output from the Oxford Nanopore MinION device. The first step in the process involved the conversion of “Fast5” in “Fastq” format. For this, we used the GPU version of the Nanopore *Guppy* basecaller (*v3*.*4*.*4*) tool (7) with the following parameters: “*flow_cell = FLO-MIN106*” and “*kit = SQK-RNA002*”.

As a second step, we performed read quality control using the *PycoQC (v2*.*5*.*0*.*21)* software (8) with standard parameters. This tool computes metrics and generates multiple quality plots for Nanopore technologies that allow the initial evaluation of the sequenced reads.

*PycoQC* can subsequently provide an overview of the overall quality of the reads. To clean the input data, we filtered the quality and read length using the *NanoFilt* (*v2*.*7*.*0)* tool (9). This tool can also analyze the sequencing summary file generated by *guppy_basecaller* to refine the filtering process. For these reasons, we filtered the reads using the sequencing summary file under the following parameters: minimum read length ≥ 500 nt and read quality ≥ 8.

Thereafter, only the filtered reads were considered in further analysis. At this point, we created an alignment process that mapped reads to a reference genome and identified the exact genomic loci corresponding to each read. Given that, we did not use primers for the amplification of the SARS-CoV-2 genome, we had to remove all the reads that were mapped with other material sequenced from the swabs. In this study, to remove the contaminating sequences, we considered humans, fungal, and bacterial reference genomes, respectively. To elaborate, we used the “Homo sapiens genome assembly *GRCh38* (*hg38*)” from the Genome Reference Consortium (10) as the human reference genome, and all sequences from both fungal and bacterial genome projects from NCBI (11). Lastly, we used the NCBI SARS-CoV-2 sequence NC_045512.2 (12) as a reference genome for SARS-CoV-2.

For each of these reference genomes, we mapped the reads using the *minimap2* (*v2*.*17-r941*) tool (13) based on earlier reports that demonstrated the effective performance of this tool with the splice-aware alignment of Nanopore Direct RNA reads against a reference genome (14). As suggested by authors who reported the performance of *minimap2*, we set the parameter k = 13 to increase the sensitivity and to map noisy Nanopore Direct RNA-seq reads.

Next, we extracted both unmapped reads and reads with mapping quality lower than 10 using the “*view*” utility in the *samtools* (*v1*.*7*) library (15) for each reference genome except for that of SARS-CoV-2.

Resultantly, we obtained a sub-set of the sequenced long-reads that did not map with the genomes of human, fungi, or bacteria.

Lastly, we mapped the remaining reads against the SARS-CoV-2 reference genome using *minimap2* tools with the same parameters used earlier. We successively analyzed the results of the mapping process using the *BCFtools* (v1.9) library (16), a set of utilities for variant calling. In particular, we first used the *mpileup* tool (16) to generate a summary of the coverage of the mapped reads on the SARS-CoV-2 reference genome at single base-pair resolution, followed by the *call* tool (17) for generating calls. We set these tools to perform as the standard consensus-caller with only the variant sites returned. The results of this pipeline were enlisted as SARS-CoV-2 variants detected in the sequenced samples from the swabs.

### Calculation of mutation frequency

#### Sanger sequencing

The samples were sequenced using the BigDye™ Terminator v3.1 Cycle Sequencing Kit (Life Technologies; Cat no 4337455) with an ABI PRISM 3100 Genetic Analyzer upgraded to the Applied Biosystems® 3130xl System (Life Technologies; Cat no 4359571). To enrich the identified mutation region in the N gene, we adopted a nested PCR approach using an outer and an inner primer sets. Both primer sets are reported in Tab. S1.

#### RT-PCR

The ten sequenced samples and the two uninfected control RNAs were retro-transcribed using the EcxcelRT™ Reverse Trascription kit (SMOBIO, Cat. No RP1300). First, 16 µL of each sample was mixed with 2 µL of 50 µM of Oligo (dT)_20_ primer and 2 µL of dNTP Mix (10 mM each). The mixes were incubated at 70 °C for 5 min and placed on ice for at least 1 min. Next, 8 µL of 5X RT Buffer (DTT), 8 µL of DEPC-Treated water, 2 µL of RNAok™ Rnase Inhibitor, and 2 µL ExcelRT™ Reverse Transcriptase was added to each sample. The reactions were incubated at 25 °C for 10 min, at 44 °C for 50 min, and at 85 °C for 5 min, and were subsequently held at 4 °C.

#### qPCR

To amplify the region containing the NC_045512:c.608_609_610delinsAAC mutation, we designed two primer sets to distinguish between a wild type (WT) and a mutated viral strain. The two sets used the same reverse primer, with divergence at the last three bases at the 3’-end of the forward primer. In fact, one forward primer was specific for the mutated sequence, whereas the other recognized the reference sequence.

We used two other sets as controls. One set consisted of the primers N2 from the 2019-Novel Coronavirus (2019-nCoV) Real-time rRT-PCR Panel (Centers for Disease Control and Prevention (CDC) Atlanta, GA 30333), which was used to confirm the presence of the N gene target (Fig. S1 and Tab. S2). The second set consisted of a custom specific primer for human *GAPDH* cDNA, which was used as an endogen control for the PCR (Fig. S2 and Tab. S3). All primer sequences are listed in Tab. S1.

Moreover, a plasmid vector carrying a synthetic reference SARS-CoV-2 N gene (Origene, Cat No: VC202563) was used as a reference control.

First, both analysis samples and uninfected control cDNA samples were diluted at a 1:2 ratio, while the reference control was concentrated to 0.8 ng/µL. Next, 4 µL of each sample in 20 µL of reaction mixture was assayed by qPCR using the Quantinova SYBR Green PCR kit (Qiagen, Cat No.: 208052) according to the manufacturer’s instructions in a Rotor-Gene Q 5plex HRM Thermocycler (Qiagen, Cat No./ID: 9001580). The following PCR thermal profile was used: 95 °C, 2 min; 95 °C, 5 s and 57 °C, 10 s for 35 cycles.

## RESULTS

To characterize the SARS-CoV-2 mutational landscape, we used RNA extracted from samples collected from five male and five female patients who tested positive for SARS-CoV-2 in the swab-test, with the corresponding Ct values ranging from 18 to 24. First, we assessed the concentrations and the fragmentation levels in the samples, as reported in the Materials and Methods. The overall samples had significantly low concentrations (in the order of pg/µL) and high fragmentation levels with an RIN index ranging from 2.6 to 2.1. Next, we prepared the pools A and B from the samples, as explained in the Materials and Methods. We prepared the former to increase the abundance of sequencing output and the latter to improve the heterogeneity in the data. We set up each pool at a final volume of 10 µL, and prepared the libraries for direct RNA-seq, as described in the Materials and Methods.

After each purification step, the concentrations were 1.2 ng/µL and 0.6 ng/µL in the initial eluates, and 0.9 ng/µL and 0.3 ng/µL in the final eluates at the pre-sequencing stage for pools A and B, respectively.

Although the library concentrations were 200 times less than that required for the protocol, we opted to load the libraries and test the suitability of MinION.

To perform a single MinION sequencing run, we loaded library A in the flow cell at the beginning of the run with 1,575 active pores enabled, while library B was loaded when 1,260 active pores remained active, as described in the Materials and Methods.

Using the *MinKNOW* and *Guppy* basecaller tools, we obtained a set of 397,465 reads in the Fastq format file.

As we stated previously, the overall quality of reads is affected by the adopted sequencing technique; i.e., the direct RNA sequencing of samples derived from the swabs. In this experiment, the average read length was lower than that of standard long-reads, and the quality of certain reads was below conventional levels. Therefore, we preferred to discard the reads that had low quality (< 8) as well as a short length (< 500 nt). We obtained a subset of 20,940 good-quality reads.

At this point, we aimed to identify and remove sequences from contaminant species contained in the swab; for this, we used the *minimap2* tool (13) to filter out reads that mapped with sequences from human, fungal, and bacterial genomes.

Using this method, we filtered 97.63% of the reads; details of the composition of these reads are provided in Tab. S4. Lastly, we attempted to map the remaining 2.37% of reads with the SARS-CoV-2 reference genome, and observed that 10.89% of the reads (i.e., 54 reads) mapped. These reads did not cover the entire SARS-CoV-2 reference genome; however, these were sufficient for mapping the N gene.

Next, we analyzed the coverage of the mapped reads and the variant calling related to N gene using SAM*tools (15)* and *BCFtools (16*) libraries. The analysis revealed the existence of the following mutation region with a quality score greater than 94.99: NC_045512:c.28881_28882_28883delinsAAC.

The N gene sequence generated is available in the GenBank database, and can be accessed at: https://www.ncbi.nlm.nih.gov/nuccore/MT457389 Fig.1 shows the results of the alignment phase with the Integrative Genomic Viewer (IGV) (*v2*.*8*.*2*) tool (17). The manner in which the reads mapped considerably well with the region of interest in the SARS-CoV-2 reference genome is apparent; also, the coordinates where the mutation appears are clearly visible.

**Fig. 1.**
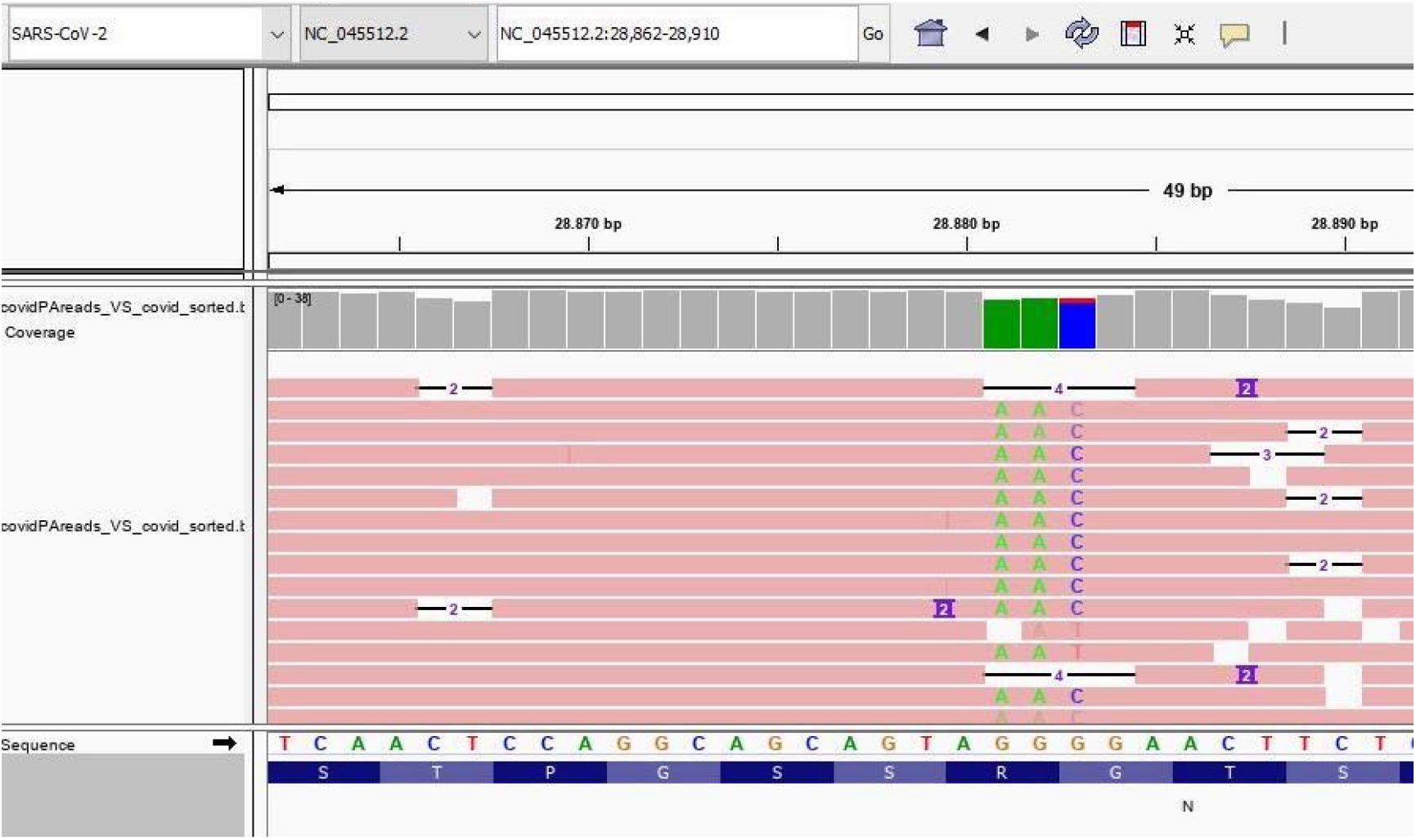
Genome IGV analysis that shows the frequency of the identified NC_045512:c.28881_28882_28883delinsAAC mutation in the reads from sequenced samples.

To determine the frequency of mutation in the samples, we adopted two different approaches: Sanger sequencing and qPCR. Using the former approach, we amplified the region that contained the mutation, as described in the Materials and Methods, prior to sequencing it. As shown in Fig. 2, all samples (100%) contained the NC_045512:c.28881_28882_28883delinsAAC mutation.

**Fig. 2.**
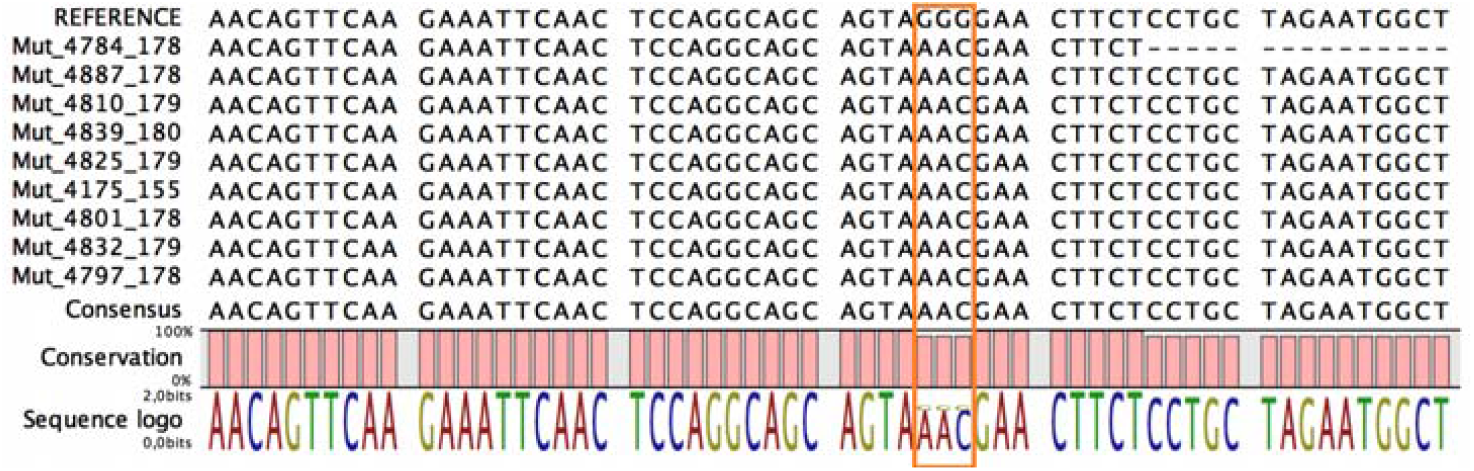
The sequences obtained by Sanger sequencing of RNA extracted from swabs who tested positive for SARS-CoV-2 infection. The rectangle covers coordinates from 28881 to 28883 for underlines as all samples contained the NC_045512:c.28881_28882_28883delinsAAC mutation.

In the qPCR assay, we used custom primer sets designed to distinguish between WT and mutated sequences, and another pair set, as reported in the Materials and Methods. We compared two uninfected controls and the synthetic SARS-CoV-2 N cDNA, which was similar to the reference control. All the samples yielded positive results with the primer set specified for the mutated sequence (Fig. S3 and Tab. S5) and negative for the WT sequence (Fig. S4 and Tab. S6), in contrast to that for the reference control, consistent with the results of the Sanger sequencing experiment. The uninfected samples tested negative for both mutated and WT sets, which confirmed the absence of a possible off-target from the host.

## DISCUSSION

Next-generation sequencing technologies can be used to detect the nucleotide sequences in analyzed samples within a short duration and at an affordable cost (18). However, these approaches depend on both the harvest and extraction protocols of the sample, which significantly influence the coverage and depth of the sequencing reads (21, 22).

Even though cDNA sequencing is considered the gold standard for the analysis of critical materials (23), DNA polymerase can introduce certain read errors during the DNA synthesis step, which can considerably affect the SNPs analysis (3, 22).

Instead, direct RNA-seq can help identify the native sequences without contamination by artefacts that are usually introduced in the in vitro amplification step, even though it requires a higher input concentration compared to cDNA sequencing methods (21) In this context, swab RNA, which has a lower concentration and is also more fragmented than in vitro RNA, may not be suitable for performing direct sequencing.

However, the performance of the MinION flow cell has improved exponentially in recent years (2); therefore, we chose to utilize MinION Mk1B to characterize the mutational landscape in the critical samples.

To maintain the sequencing input concentration and its heterogeneity, we combined RNA samples from two pools (A and B). At the end of the library workflow, both samples were present at concentrations that were approximately 200 times less than that required for the protocol. In fact, owing to insufficient starting material, we observed that only a limited number of pores were active at a time, as shown in the MinKNOW duty time plot in Fig. 3.

**Fig. 3.**
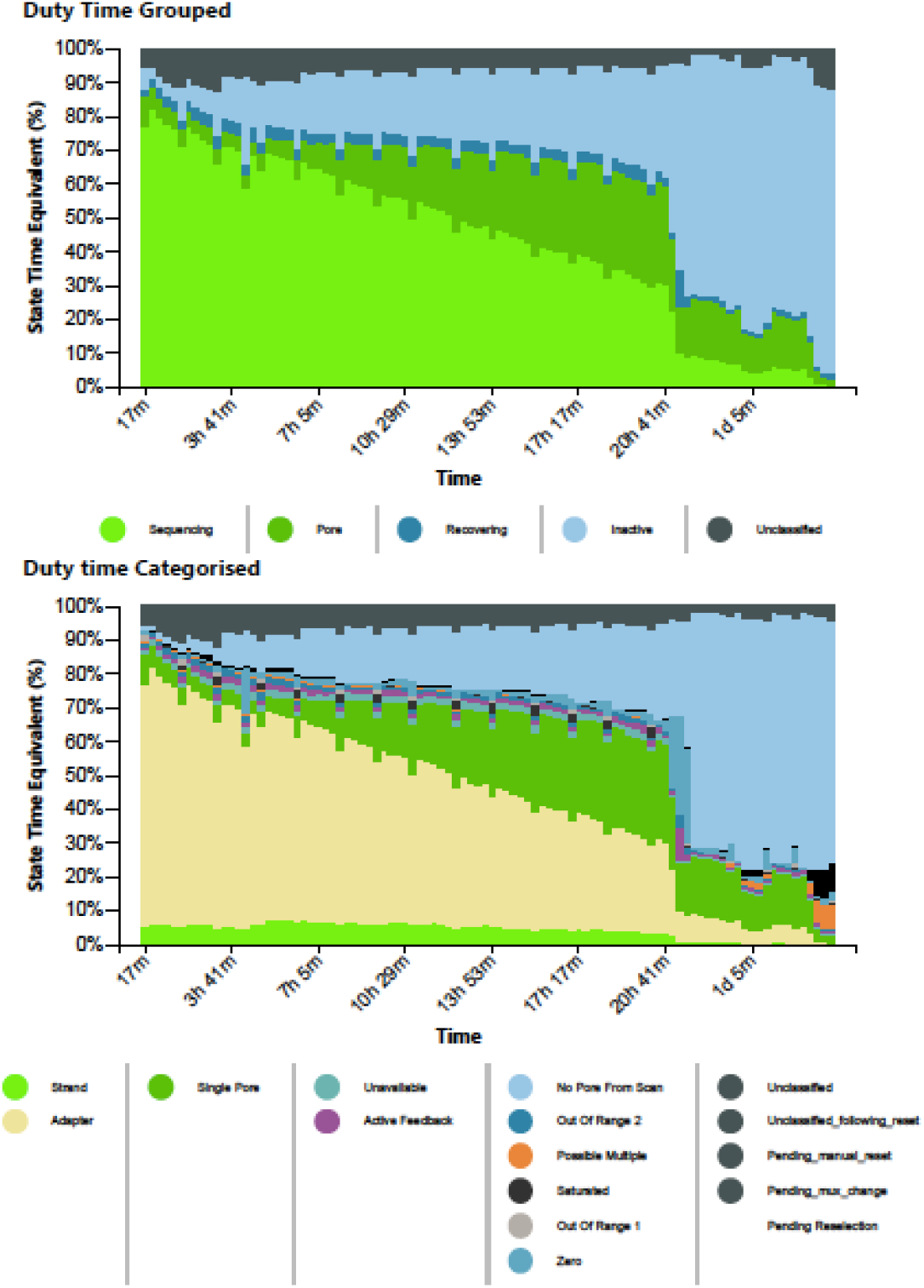
The MinKNOW duty time plots depict the sum of total channel activity within a particular period of time. The number of sequencing pores decrease over time. In the categorized plot, the imbalance of adapters is apparent.

Resultantly, we only obtained a few reads (approximately 0.3%), which showed significant variant calling. Next, we investigated whether findings from other studies on SARS-CoV-2 were consistent with our findings. To this end, we considered sequence variations from the China National Center for Bioinformation (CNCB) portal (23). CNCB is the largest, daily-updated, publicly available SARS-CoV-2 genome variation repository. It contains more than 21,000 high-quality, human-hosted, complete SARS-CoV-2 genomes (last download on 12-Jun-2020) from several countries worldwide. We retrieved this dataset and prepared a script to check for the presence of the identified mutation region (NC_045512:c.28881_28882_28883delinsAAC) in the sequences of N gene present in CNCB’s dataset. As shown in Fig. 4A, we considered three sets: 1) cases that only contained identified mutations, 2) cases that also contained identified mutations, and 3) cases that did not contain the identified mutations. We found that the same N gene sequence reported by us was reported in approximately 18% of COVID-19 cases, and we took into account the distribution of these cases over time.

**Fig. 4.**
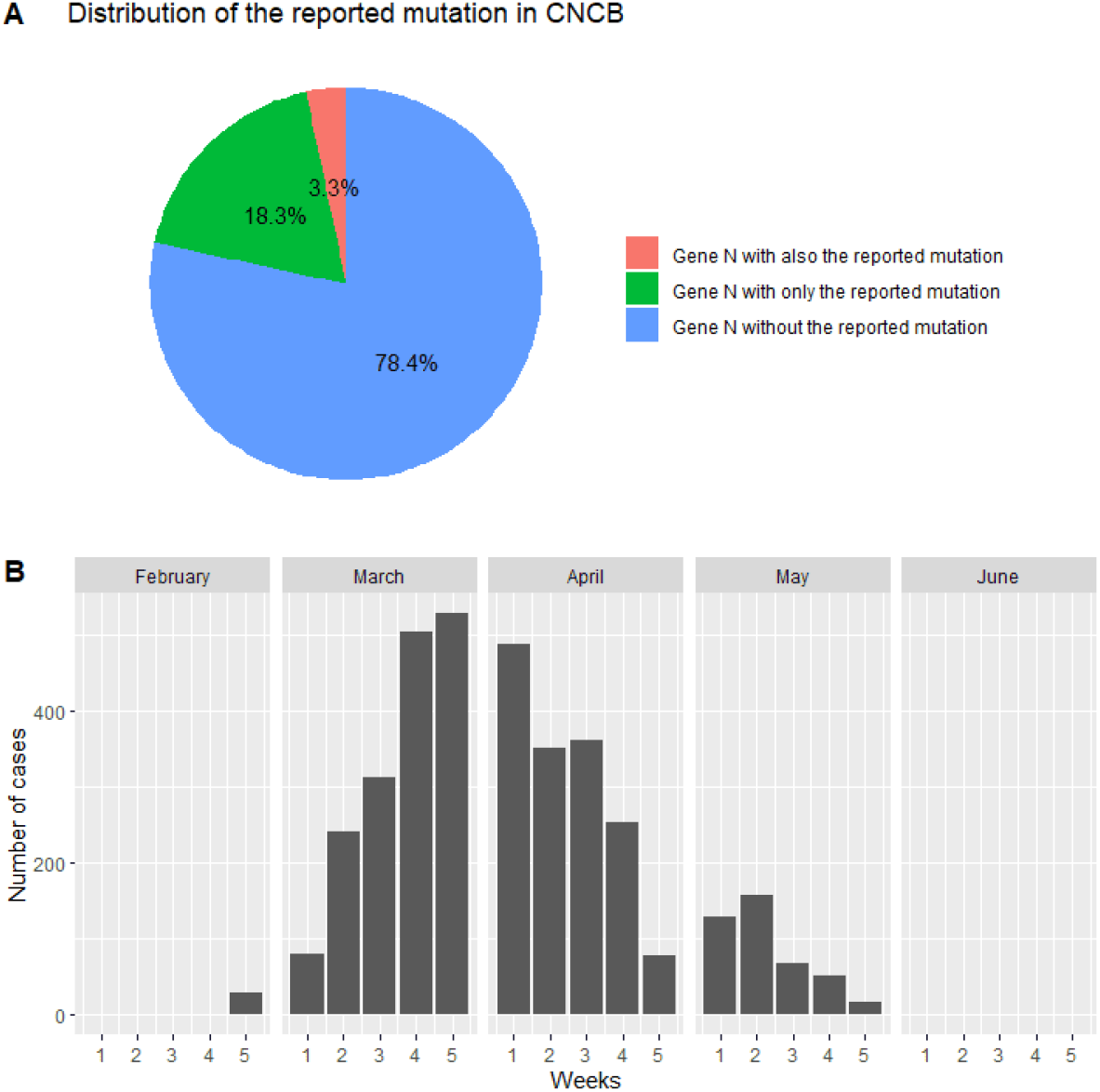
Identification of the reported mutation in the CNCB database. (A) Strains isolated from 18.3% of COVID-19 cases (over 21,000 complete genomes sequenced from isolated strains) contained the reported mutation. (B) Plot depicting the distribution of these cases from the end of February to the beginning of June.

As shown in Fig. 4B, we reported the presence of the mutation in cases treated per week from February to June, and noticed that the majority of cases were reported between the second week of March and the fourth week of April. This period coincided with that of swab collection from patients.

Lastly, we wanted to compare the in silico results using Sanger sequencing and qPCR techniques, as described in the Results section. The presence of the NC_045512:c.28881_28882_28883delinsAAC mutation was confirmed using both methods in all the samples used in this study. Our results validated the approach of direct RNA-seq with Oxford Nanopore MinION technology. Additionally, the presence of molecular targets for the identification of specific genomic mutations was confirmed, and could be considered advantageous for both surveillance and epidemiologic studies.

## Data Availability

The undersigned Davide Vacca and Antonino Fiannaca as first coauthors of the titled manuscript "Direct RNA nanopore sequencing of SARS-CoV-2 extracted from critical material from swabs", declare that all data referred to this work are aviable for scientific aimes.

https://www.ncbi.nlm.nih.gov/nuccore/MT457389

## Data availability

The data that support the findings of this study are available from the corresponding author upon reasonable request.

Supplemental material is available to link: https://www.dropbox.com/sh/yphu9kzd64mnrl7/AAC8khoPjZ8wSeZRk_6DFd6a?dl=0

## Contributions

DV: Data curation, Formal analysis, Methodology, Software, Writing, Editing; AF: Data curation, Formal analysis, Methodology, Software, Writing, Editing; FT: Investigation, Methodology, Resources, Visualization; VC: Investigation, Methodology, Visualization; LLP: Visualization, Review, Editing; WM: Investigation, Resources, Visualization; AG: Data curation, Visualization; MLR: Data curation, Methodology, Visualization, Review, Editing; CM: Data curation, Methodology, Resources, Visualization; GM: Data curation, Methodology, Visualization; BB: Data curation, Visualization; AC: Data curation, Investigation, Resources, Visualization; RM: Resources, Visualization; GI: Resources, Visualization; FV: Conceptualization, Data curation, Investigation, Supervision, Visualization, Writing, Review, Editing; CT: Conceptualization, Data curation, Funding acquisition, Investigation, Project administration, Supervision, Visualization, Writing, Review, Editing; AU: Conceptualization, Data curation, Investigation, Supervision, Visualization, Writing, Review, Editing

## Conflicts of interest

All authors no reported conflicts.

## Notes

### Competing Interest Statement

The authors have declared no competing interest.

### Funding Statement

This work was supported by AIRC (Associazione Italiana per la Ricerca sul Cancro)

### Author Declarations

EC N˚11/18DEC2020 - University of Palermo - Department of Sciences for Health Promotion and Mother-Child Care G. D Alessandro - Piazza delle cliniche N˚2 - zip code 90127 -Palermo - Italy.

