## Supplemental_Figures_and_Tables for "Direct RNA nanopore sequencing of SARS-CoV-2 extracted from critical material from swabs"

### Supplementary Figures

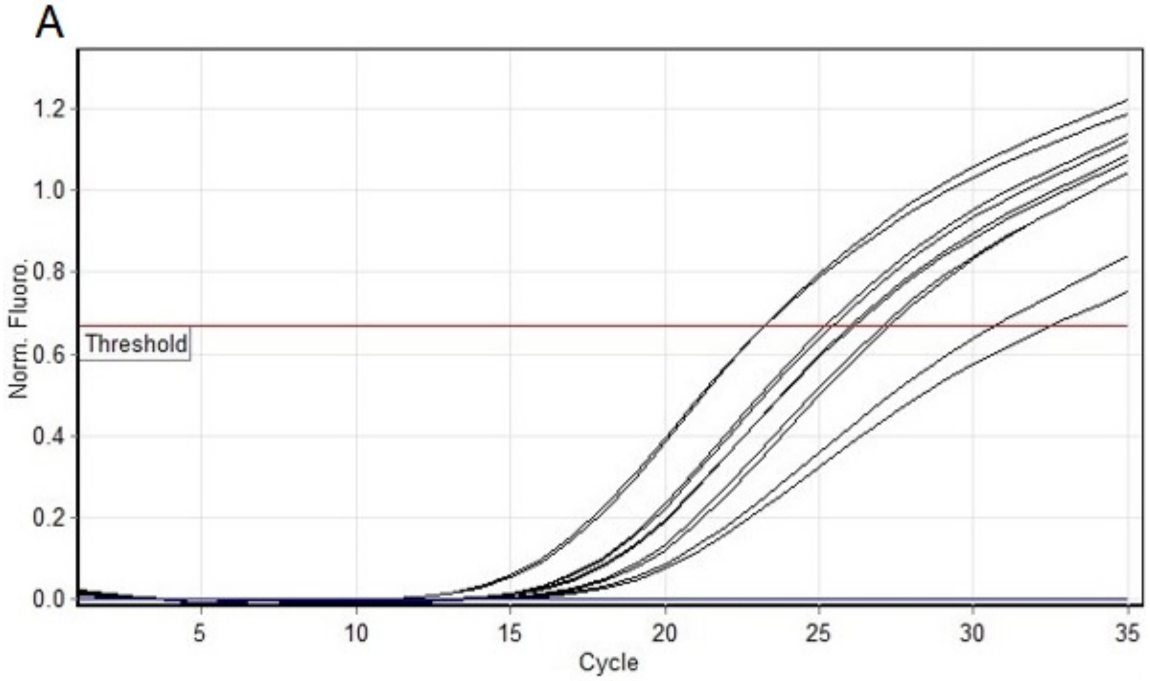


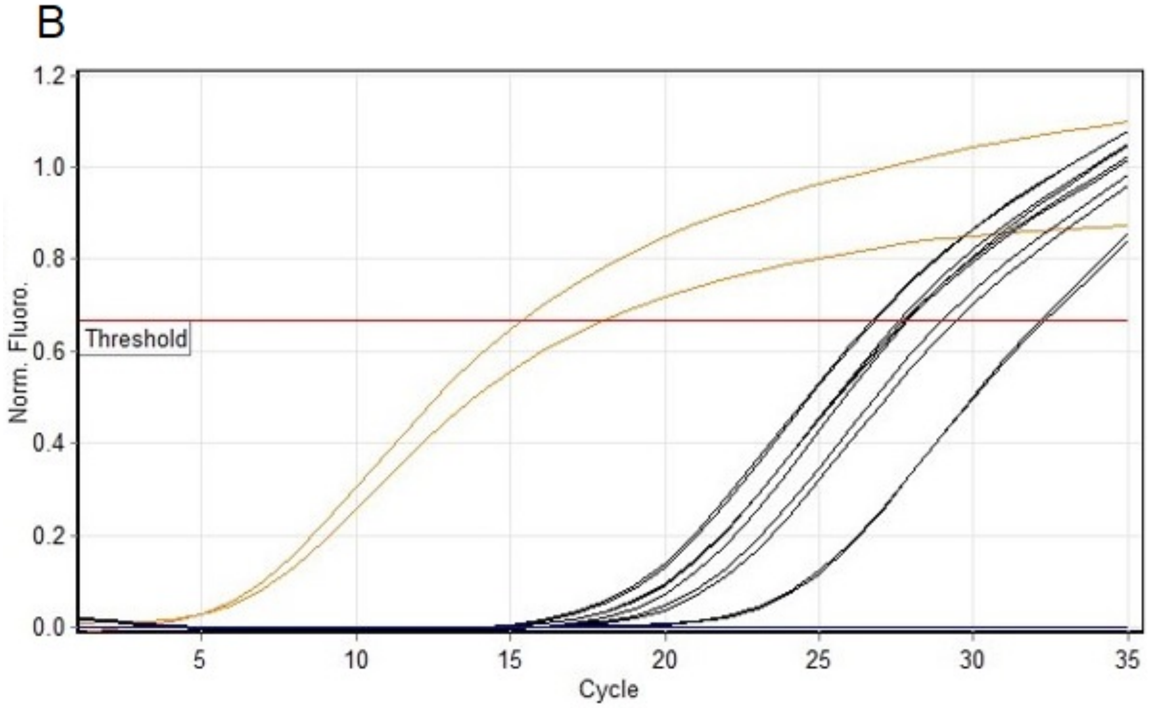


**Fig. S1:** Amplification curves in the qPCR assay with primer set N2 from 2019-Novel Coronavirus (2019-nCoV) Real-time rRT-PCR Panel, from IG1 to IG5 (A) and from IG6 to IG10 with negative and positive control samples (B), as reported in the Materials and Methods. The IG and N reference samples tested positive in the assay compared to the brain samples. Relative Ct values are reported in Tab. S3.

A

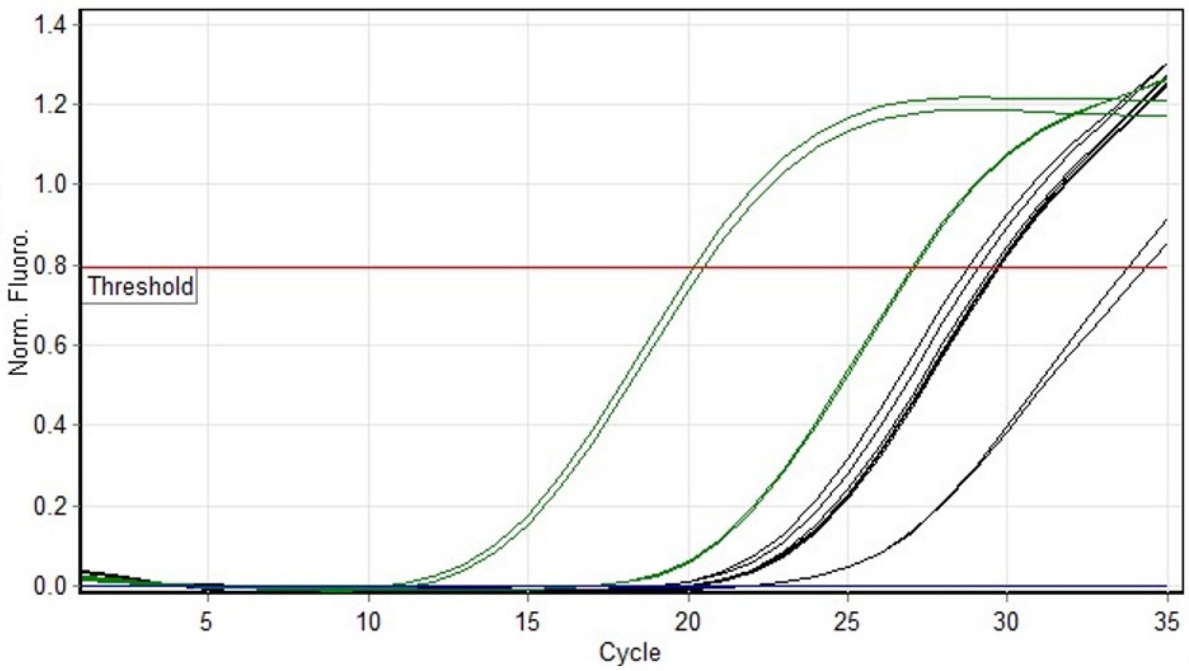


B

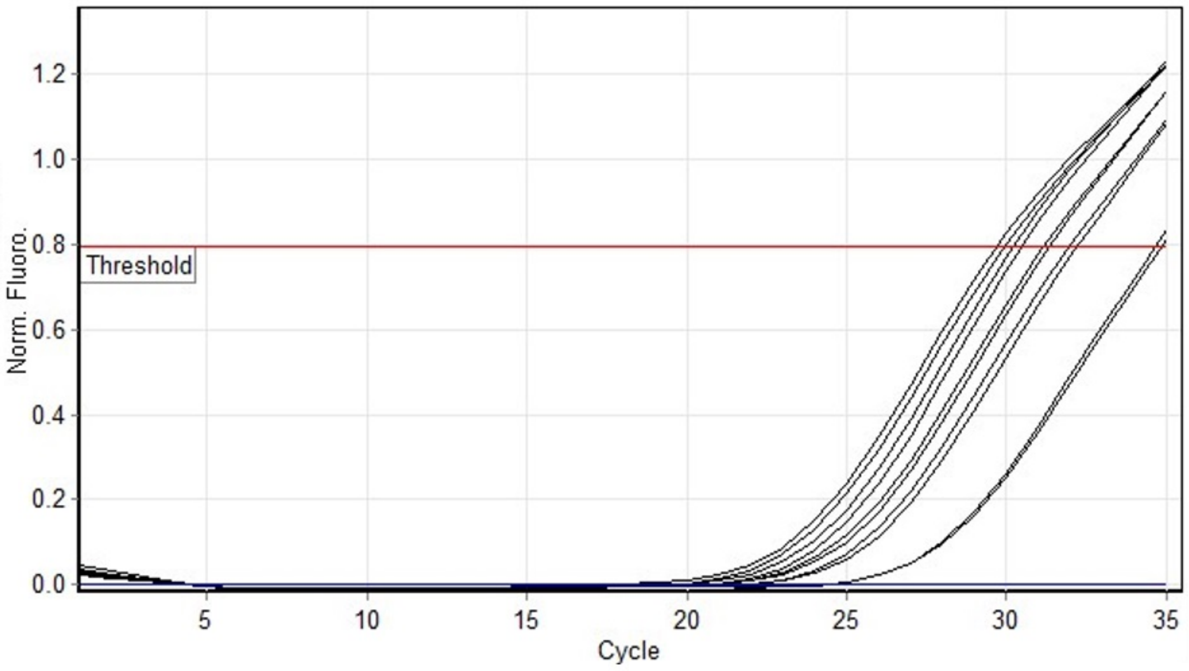


**Fig. S2:** Amplification curves in the qPCR assay with primer set specific for Human housekeeping GAPDH, from IG1 to IG5 (A) and from IG6 to IG10 with negative and positive control samples (B), as reported in the Materials and Methods. The IG and brain samples tested positive in the assay compared to the synthetic SARS-CoV-2 N gene. Relative Ct values are reported in Tab. S4.

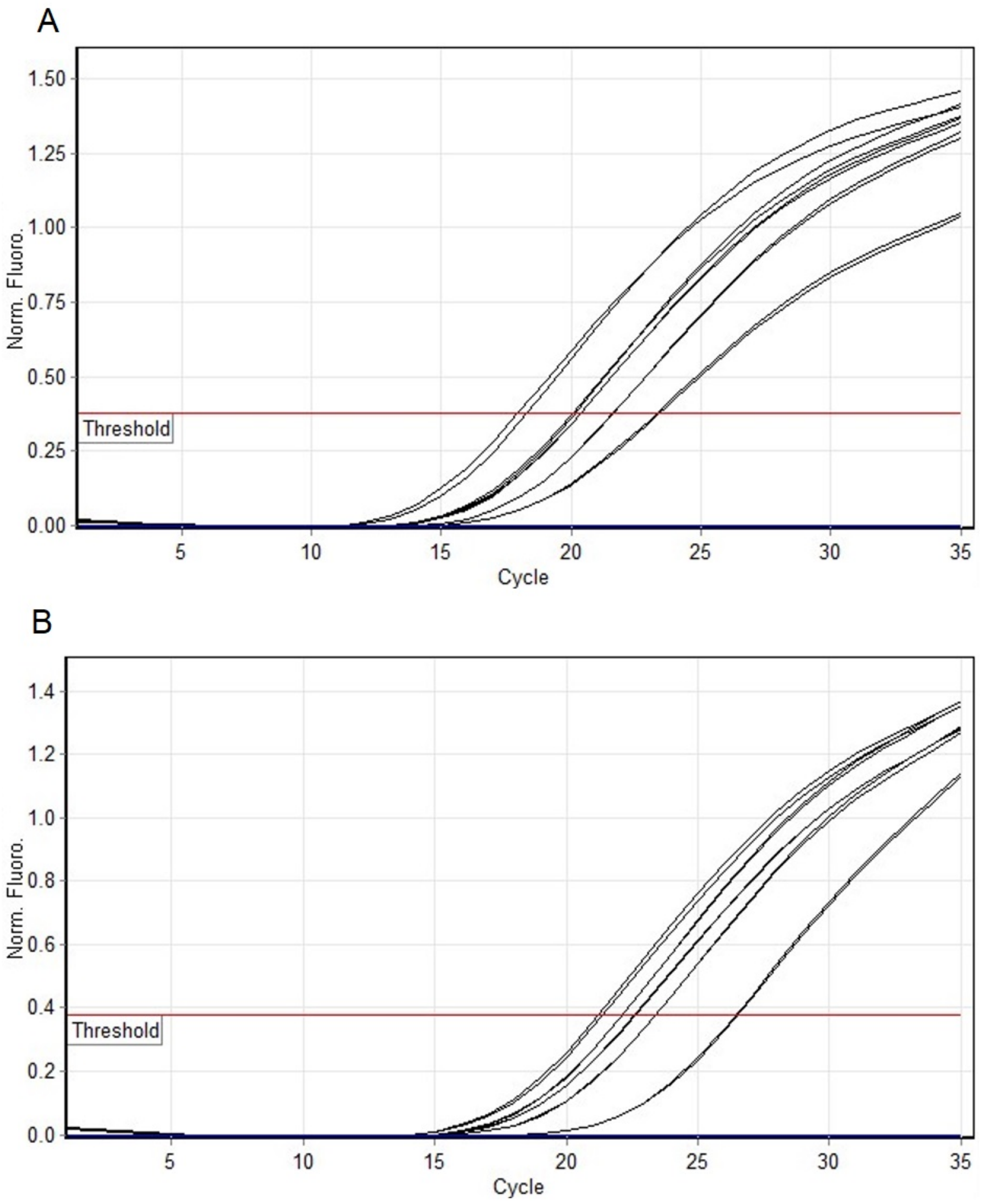


**Fig. S3:** Amplification curve in the qPCR assay with the primer set specific for the mutated region, from IG1 to IG5 (A) and from IG6 to IG10 with negative and positive control samples (B), as reported in the Materials and Methods. All the IG samples exhibited mutation (frequency of 100%) compared to the reference and brain biopsy samples. Relative Ct values are reported in Tab. S5.

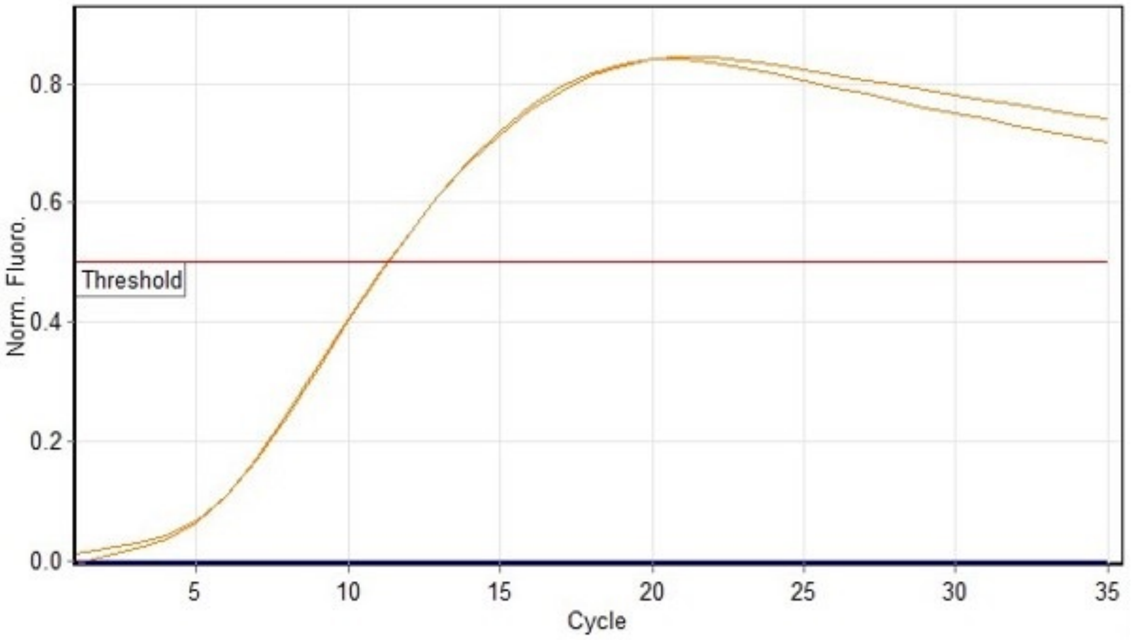


**Fig. S4:** Amplification curves in the qPCR assay for samples using the WT primer set. The plasmid vector carrying a synthetic reference SARS-CoV-2 N gene was the unique sample positive compared to other samples, as reported in the Materials and Methods. Relative Ct values are reported in Tab. S6.

### Supplementary Tables

**Tab. S1:** The primer sequences of the sets used for the enrichment of the mutated region in the Sanger sequencing workflow, and of the primers specific for the analyzed targets in the real-time PCR experiment, as reported in the Materials and Methods.

| **Primer set name** | **Primer sequence (5' → 3')** | **Size (bp)** |
|---|---|---|
| Sanger_nCoV_Gene N_OUT | F: CAAGGCGTTCCAATTAACA<br>R: CAGCAGGAAGAAGAGTCA | 985 |
| Sanger_nCoV_Gene N_INN | F: TACGCAGAAGGGAGCAGAG<br>R: AGCAGCAAAGCAAGAGCA | 155 |
| 2019-nCoV_N2 | F: TTACAAACATTGGCCGCAAA<br>R: GCGCGACATTCCGAAGAA | 67 |
| 2019-nCoV_28881_28882_28883-N | F (wt): AACTCCAGGCAGCAGTAGGG<br>F (mut): AACTCCAGGCAGCAGTAAAC<br>R: TTGGCCTTGTTGTTGTTGGC | 142 |
| GAPDH (NM_002046.7) | F: GATTTGGTCGTATTGGGCGC<br>R: CATGTAAACCATGTAGTTGAGGTCA | 110 |

**Tab. S2:** CT values in the qPCR assay for samples derived from: swabs collected from ten SARS-CoV-2-positive patients (from IG1 to IG10); brain biopsy (n7 and n9); synthetic reference N gene and No template controls (ntc), with primer set N2 from 2019-Novel Coronavirus (2019-nCoV) Real-time rRT-PCR Panel, as reported in the Materials and Methods

| CT Values – Threshold 0.66697 | | | |
|---|---|---|---|
| Color | Name | Type | Ct |
| | ig1 | Unknown | 32.49 |
| | ig1 | Unknown | 30.65 |
| | ig2 | Unknown | 23.23 |
| | ig2 | Unknown | 23.22 |
| | ig3 | Unknown | 25.38 |
| | ig3 | Unknown | 25.17 |
| | ig4 | Unknown | 25.97 |
| | ig4 | Unknown | 26.07 |
| | ig5 | Unknown | 27.02 |
| | ig5 | Unknown | 27.24 |
| | n7 | Unknown | |
| | n7 | Unknown | |
| | n9 | Unknown | |
| | n9 | Unknown | |
| orange | N reference | Unknown | 17.95 |
| orange | N reference | Unknown | 15.38 |
| | ig6 | Unknown | 27.89 |
| | ig6 | Unknown | 27.62 |
| | ig7 | Unknown | 26.79 |
| | ig7 | Unknown | 26.73 |
| | ig8 | Unknown | 32.32 |
| | ig8 | Unknown | 32.16 |
| | ig9 | Unknown | 27.84 |
| | ig9 | Unknown | 27.76 |
| | ig10 | Unknown | 29.01 |
| | ig10 | Unknown | 29.45 |
| blue | ntc | Unknown | |
| blue | ntc | Unknown | |

**Tab. S3**: CT values of the qPCR assay for samples derived from: swabs collected from ten SARS-CoV-2-positive patients (from IG1 to IG10); brain biopsy (n7 and n9); synthetic reference N gene and No template controls (ntc), with primer set specific for Human housekeeping *GAPDH*, as reported in the Materials and Methods.

| CT Values – Threshold 0.79373 | | | |
|---|---|---|---|
| Color | Name | Type | Ct |
| | ig1 | Unknown | 34.33 |
| | ig1 | Unknown | 33.77 |
| | ig2 | Unknown | 29.66 |
| | ig2 | Unknown | 29.55 |
| | ig3 | Unknown | 29.73 |
| | ig3 | Unknown | 29.77 |
| | ig4 | Unknown | 28.79 |
| | ig4 | Unknown | 29.11 |
| | ig5 | Unknown | 29.68 |
| | ig5 | Unknown | 29.71 |
| green | n7 | Unknown | 20.15 |
| green | n7 | Unknown | 20.49 |
| green | n9 | Unknown | 27.08 |
| green | n9 | Unknown | 27 |
| | N reference | Unknown | |
| | N reference | Unknown | |
| | ig6 | Unknown | 30.23 |
| | ig6 | Unknown | 30.48 |
| | ig7 | Unknown | 31.16 |
| | ig7 | Unknown | 31.33 |
| | ig8 | Unknown | 34.81 |
| | ig8 | Unknown | 34.63 |
| | ig9 | Unknown | 29.71 |
| | ig9 | Unknown | 29.96 |
| | ig10 | Unknown | 31.95 |
| | ig10 | Unknown | 32.19 |
| blue | ntc | Unknown | |
| blue | ntc | Unknown | |

**Tab. S4**: Result of the reads alignment process given by the bioinformatics pipeline.

| | Number of Reads | Percentage of good-quality reads |
|---|---|---|
| Reads from MinION sequencing technlogy | 397465 | |
| Good-quality reads (quality > 8, length > 500nt) | 20940 | 100% |
| Fungi (Saccharomyces cerevisiae) | 20351 | 97.19% |
| Homo Sapiens (Chromosome MT) | 93 | 0.44% |
| SARS-CoV-2 | 54 | 0.26% |
| Bacteria | 0 | 0% |
| ND | 442 | 2.11% |

**Tab. S5:** Ct values in the qPCR assay for samples derived from: swabs collected from ten SARS-CoV-2-positive patients (from IG1 to IG10); brain biopsy (n7 and n9); synthetic reference N gene and No template controls (ntc), with the primer set specific for the mutated region.

| CT Values – Threshold 0.37927 | | | |
|---|---|---|---|
| Color | Name | Type | Ct |
| | ig1 | Unknown | 23.31 |
| | ig1 | Unknown | 23.43 |
| | ig2 | Unknown | 18.34 |
| | ig2 | Unknown | 17.97 |
| | ig3 | Unknown | 20.16 |
| | ig3 | Unknown | 20.08 |
| | ig4 | Unknown | 20.38 |
| | ig4 | Unknown | 20.37 |
| | ig5 | Unknown | 21.65 |
| | ig5 | Unknown | 21.65 |
| | n7 | Unknown | |
| | n7 | Unknown | |
| | n9 | Unknown | |
| | n9 | Unknown | |
| | N reference | Unknown | |
| | N reference | Unknown | |
| | ig6 | Unknown | 22.18 |
| | ig6 | Unknown | 22.18 |
| | Ig7 | Unknown | 21.44 |
| | Ig7 | Unknown | 21.26 |
| | ig8 | Unknown | 26.46 |
| | ig8 | Unknown | 26.54 |
| | ig9 | Unknown | 22.67 |
| | ig9 | Unknown | 22.63 |
| | ig10 | Unknown | 23.42 |
| | ig10 | Unknown | 23.43 |
| blue | ntc | Unknown | |
| blue | ntc | Unknown | |

**Tab. S6:** : Ct values in the qPCR assay for samples derived from: swabs collected from ten SARS-CoV-2-positive patients (from ig1 to ig10); brain biopsy (n7 and n9); synthetic reference N gene and No template controls (ntc), using the WT primer set.

| CT Values – Threshold 0.50197 | | | |
|---|---|---|---|
| Color | Name | Type | Ct |
| | ig1 | Unknown | |
| | ig1 | Unknown | |
| | ig2 | Unknown | |
| | ig2 | Unknown | |
| | ig3 | Unknown | |
| | ig3 | Unknown | |
| | ig4 | Unknown | |
| | ig4 | Unknown | |
| | ig5 | Unknown | |
| | ig5 | Unknown | |
| | n7 | Unknown | |
| | n7 | Unknown | |
| | n9 | Unknown | |
| | n9 | Unknown | |
| orange | N reference | Unknown | 11.28 |
| orange | N reference | Unknown | 11.30 |
| | ig6 | Unknown | |
| | ig6 | Unknown | |
| | ig7 | Unknown | |
| | ig7 | Unknown | |
| | ig8 | Unknown | |
| | ig8 | Unknown | |
| | ig9 | Unknown | |
| | ig9 | Unknown | |
| | ig10 | Unknown | |
| | ig10 | Unknown | |
| blue | ntc | Unknown | |
| blue | ntc | Unknown | |
